# Cardiac Troponin T Elevation Predicts Mortality in Hospitalized COVID-19 Patients

**DOI:** 10.1101/2025.07.03.25330855

**Authors:** Katelyn A. Bruno, R. Scott Wright, Joshua Culberson, Mikolaj A. Wieczorek, David O. Hodge, Patrick W. Johnson, Yomary A. Jimenez, Emily R. Whelan, Jose M. Malavet, Kathryn F. Larson, Jonathon W. Senefeld, Chad C. Wiggins, Stephen A. Klassen, Jacob Ricci, Taimur Sher, Rickey E. Carter, Michael J. Joyner, DeLisa Fairweather, Allan S. Jaffe

## Abstract

**Objective:** To evaluate if cardiac troponin values predict poor outcomes in COVID-19 patients across the range of patients of different sex and age.

**Methods:** We examined high-sensitivity cardiac troponin T (hs-cTnT) levels in 1,050 severely ill hospitalized COVID-19 patients who had hs-cTnT data available and participated in the Expanded Access Program for convalescent plasma study during the first wave (April-August 2020) of the COVID-19 pandemic.

**Results:** We observed a continuous relationship between hs-cTnT levels and mortality in hospitalized males and females with COVID-19. This finding was present regardless of sex or age.

**Conclusion:** These data indicate the prognostic ability of hs-cTnT to predict mortality in hospitalized COVID-19 patients across all relevant patient groups.

**Clinical Trials registration number:** NCT04338360

**Clinical Perspective:**

- This study evaluated the prognostic value of elevated hs-cTnT in patients hospitalized with COVID-19 who received convalescent plasma.
- There was a continuous relationship between elevated hs-cTnT values and mortality in both males and females hospitalized for COVID-19.
- The magnitude of the prognostic value of elevated hs-cTnT varied by sex and age, suggesting that these covariates are important to consider in this population.

## Introduction

Cardiac troponin values—including cardiac troponin I (cTnI) and T (cTnT) proteins in the troponin complex — represent highly specific and sensitive markers of myocardial injury and are widely used in clinical practice to diagnose acute coronary syndromes and assess general cardiovascular risk profiles.^1^ Prior studies have shown that increases in cardiac troponin (cTn) are associated with poor outcomes in patients with COVID-19.^2-9^ In several studies, low circulating cTnT levels (<6ng/L) were associated with a good prognosis,^4,5^ as previously reviewed.^9^ However, many of the studies evaluated cTnI (and not cTnT), used small or modest-sized populations, did not utilize high-sensitivity cTn (hs-cTn) assays, or did not evaluate sex-specific metrics.^2,3,6-8^ To address these limitations, this study leveraged hs-cTnT assay data from the US COVID Convalescent Plasma Expanded Access Program (EAP) Trial to investigate the ability of cTnT levels to predict mortality using several well established cut-off thresholds according to sex and age in a large number of patients.

The EAP enrolled 105,717 patients in a trial to examine the safety and effectiveness of convalescent plasma treatment on patients hospitalized with moderate to severe COVID-19.^10,11^ Cardiac cTn values were obtained as part of the initial enrollment information. However, data on cTn values were not provided for many patients in the study because healthcare services were often overwhelmed. Nevertheless, we were able to obtain hs-cTnT levels for 1,231 patients in the EAP. In this study we evaluated whether there were differences in the relationship of hs-cTnT levels to predict mortality using a variety of widely accepted cut-off thresholds in females vs. males and younger vs. older patients with COVID-19. We also determined whether low hs-cTnT values were predictive of a good outcome in this population, as has been previously reported.^4,5^ The high number of critically ill COVID-19 patients examined during the EAP provided a unique opportunity to investigate these questions.

## Methods

### Ethics Statement

Data used for this study received approval from the Mayo Clinic Institutional Review Board (IRB) (Mayo Clinic IRB#: 20-003312 and IRB#: 21-004150). Written informed consent was obtained from patients or their legally authorized representatives. Mayo Clinic served as the academic research organization coordinating the US study and provided regulatory oversight for all sites and investigators (Clinical Trials Registration Number: NCT04338360).^11^ A Data and Safety Monitoring Board oversaw the safety analyses and advised the regulatory sponsor (i.e., US FDA) and the Mayo Clinic IRB on risk.^10^ Study data were deposited with the US FDA. This study conformed to the principles outlined in the Declaration of Helsinki.

### Expanded Access Program

As previously described,^10,11^ the US COVID Convalescent Plasma Expanded Access Program (EAP) was a pragmatic clinical trial (NCT04338360) designed to determine whether 1) convalescent plasma was safe^10^ and 2) whether treatment with convalescent plasma reduced mortality^12^ in patients hospitalized with severe or life-threatening COVID-19. The study utilized data collected from adult (≥18 years of age) hospitalized patients with severe or life-threatening COVID-19 at the beginning of the pandemic (April-August 2020).^11^ Severe COVID-19 was defined as dyspnea, respiratory frequency ≥30/min, blood saturation ≤93%, partial pressure of arterial oxygen to fraction of inspired oxygen ratio <300, and lung infiltrates >50% within 24 to 48 hours of hospital admission. Life-threatening COVID-19 was defined as one or more of the following: respiratory failure, septic shock, and/or multiple organ dysfunction/failure. All data used in this study was from patients treated with convalescent plasma during their hospitalization.

### Data

Information on patient demographics, medical history, laboratory values including cTnT, and mortality were obtained from case report forms (Mayo Clinic IRB#: 20-003312). Data on gender was collected on the case report form, but only female and male patients are included in this analysis due to the small number of patients reported for ‘other’ gender categories. We received approval to obtain additional information (sent out as a survey) at the hospital level regarding specific troponin assay and the instruments that were used for the assays (Mayo Clinic IRB#: 21-004150). This included information on the instrument and manufacturer, the specific cTnT assay that was used, and any cut-offs for the assay.

### Troponin Assays

Many different hs-cTnT assays and assay systems (i.e., instruments) exist.^13,14^ The lowest value for hscTnT that is allowed to be reported in the US according to the FDA is a value of <6ng/L, which is the limit of quantitation (LOQ). The LOQ is the lowest concentration of an analyte that may be reliably measured with acceptable precision and accuracy. The FDA approved blood sample type is lithium heparin. The assay’s limit of detection (LOD) is 5ng/L, depending on the instrument used. Sex-specific values for hs-cTnT are ≤10ng/L in females and ≤15ng/L in males, based on prior normal range data.^13,15^ The European Society of Cardiology (ESC) recommends a combined male and female threshold cut-off value of 14ng/L.^16^ Troponin T levels used in this analysis were evaluated according to all of these potential cut-off values: FDA lowest cut-off value of 6ng/L, ESC combined male and female cut-off value of 14ng/L, and the sex-specific cutoff value of ≤10ng/L for females and ≤15ng/L for males. Additionally, raw data for cTnT values were also analyzed as a continuous variable. Hs-cTnT values were provided by hospital sites at any time from the time of enrollment in the EAP through completion of the study. Additionally, patients may have been enrolled in the EAP at any time during their hospital stay. Furthermore, medications given during the hospital stay may have been administered prior to the hs-cTnT blood sample being drawn.

### Statistical Analyses

Baseline study characteristics were compared between patients with normal versus elevated (i.e., abnormal) hs-cTnT levels. Continuous variables were summarized using the mean and standard deviation (SD) and compared using the two-sample t-test for unequal variances. Categorical variables were reported and compared using the Chi-square two-sample test, including the number and percentage of patients.

Associations between hs-cTnT levels and 30-day mortality were evaluated using logistic regression models, with odds ratios (ORs) and 95% confidence intervals (CIs) reported. Data were adjusted for pre-existing cardiovascular disease (CVD) due to the potential influence of CVD on troponin levels.^17^ Troponin levels were analyzed both as a continuous variable and across different categorical cutoff levels based on various clinical cut-off thresholds (i.e., FDA 6ng/L, ESC 14 ng/L, and sex-specific cut-off values of ≤10ng/L for females and ≤15ng/L for males). Each troponin variable was used as a predictor in univariate models and multivariate models, the latter of which adjusted for the presence of pre-existing cardiovascular conditions. For the logistic regression models involving hs-cTnT as a continuous variable, the predictor was transformed to a logarithmic scale. This analysis was employed for the overall cohort as well as for subgroups based on age (under 65 years, over 65 years) and sex (males, females). To test whether the association between hs-cTnT and mortality differed by sex, a multiplicative interaction term between log-transformed troponin and sex was included in a separate logistic regression model. All analyses were performed using R Statistical Software (R Foundation for Statistical Computing, Vienna, Austria)^11^, p-values less than 0.05 were considered statistically significant, and no adjustments were made for multiple comparisons.

## Results

### Patient groups

The US COVID-19 Convalescent Plasma EAP enrolled 105,718 patients via 12,576 physicians at 2,247 hospitals during the early COVID-19 pandemic (April-August 2020).^11^ In this study, a total of 1,231 patients reported hs-cTnT levels on the case report forms. Of these, 741 were male (60.2%), and 490 were female (39.8%). There were 610 patients under the age of 65 (49.6%) and 621 patients over the age of 65 (50.4%). In some cases, the data was reported on the case report form as being above or below a certain cut-off value rather than providing the actual value. In the subgroup of 1,050 patients that had numerical values available, 643 were male (61.2%) and 407 were female (38.8%). In this subgroup, there were 498 patients under age 65 (47.4%) and 552 patients over age 65 (52.6%). Thus, the two patient groups were similar.

### Demographic characteristics of patients with sex-specific elevated hs-cTnT levels

To evaluate the effect of elevated hs-cTnT on demographic characteristics we used the sex-specific hs-cTnT values of ≤10ng/L for females and ≤15ng/L for males (**Table 1**). Compared to patients with normal hs-cTnT levels, patients with elevated hs-cTnT levels were older at enrollment (60 vs. 66 years of age, *P*<0.001), less obese (BMI 32.8 vs. 31.5 kg/m^2^, *P*<0.014), had severe or life-threatening COVID-19 conditions (62.0% vs. 79.0%, *P*<0.001), a higher incidence of oxygen desaturation ≤93% (43.7% vs. 51.6%, *P*=0.014), and a greater presence of respiratory failure (42.7% vs. 50.5%, *P*=0.037). They were also transfused with convalescent plasma later after hospitalization (6.8 vs. 7.7 days, *P*=0.019), with day 0 being the first day of their hospital admission. Additionally, they received less Remdesivir (49.4% vs. 29.5%, *P*<0.001) but more steroids (57.4% vs. 66.0%, *P*=0.013) and Hydroxychloroquine (16.0% vs. 32.2%, *P*<0.001).

**Table 1:**
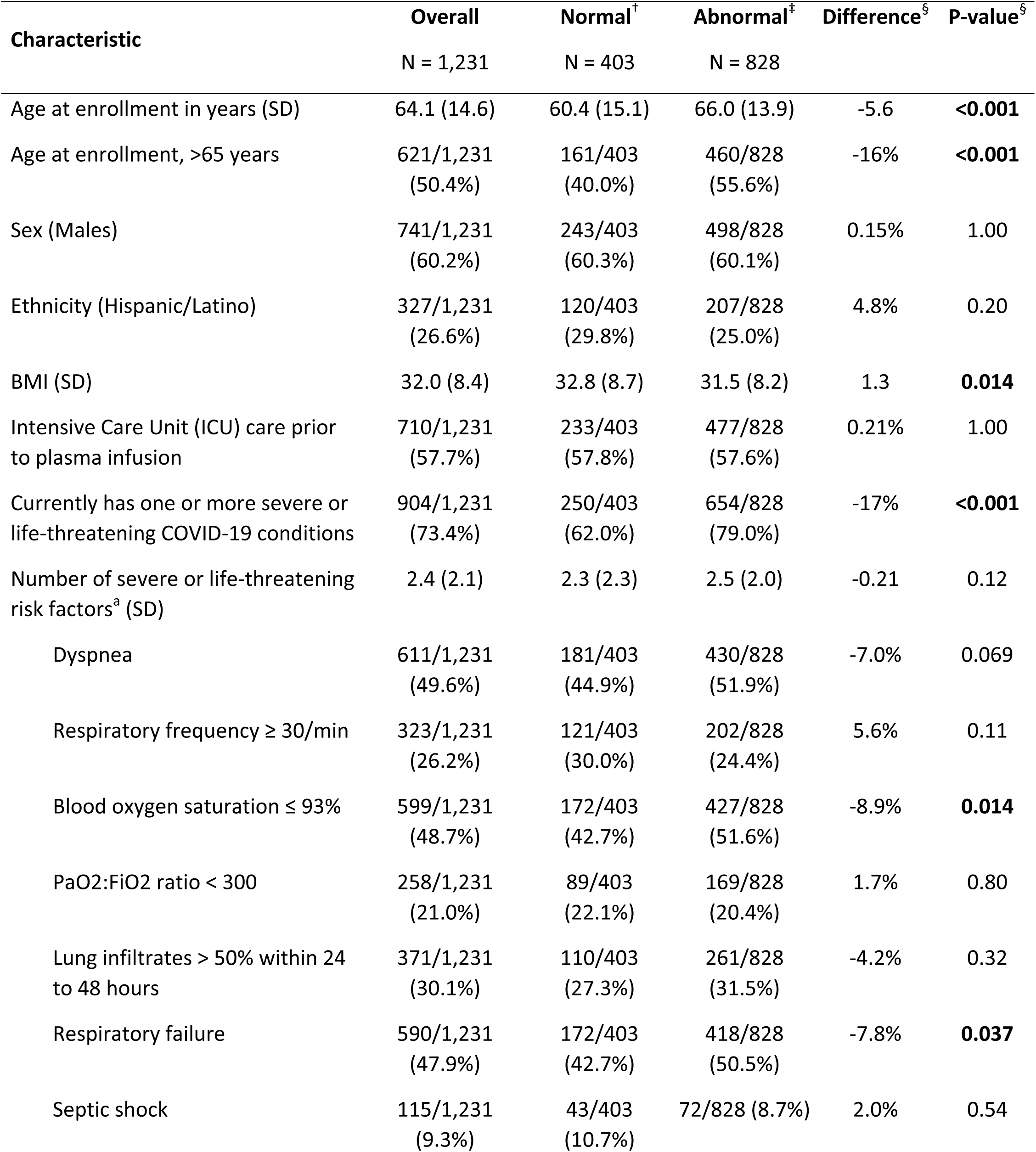
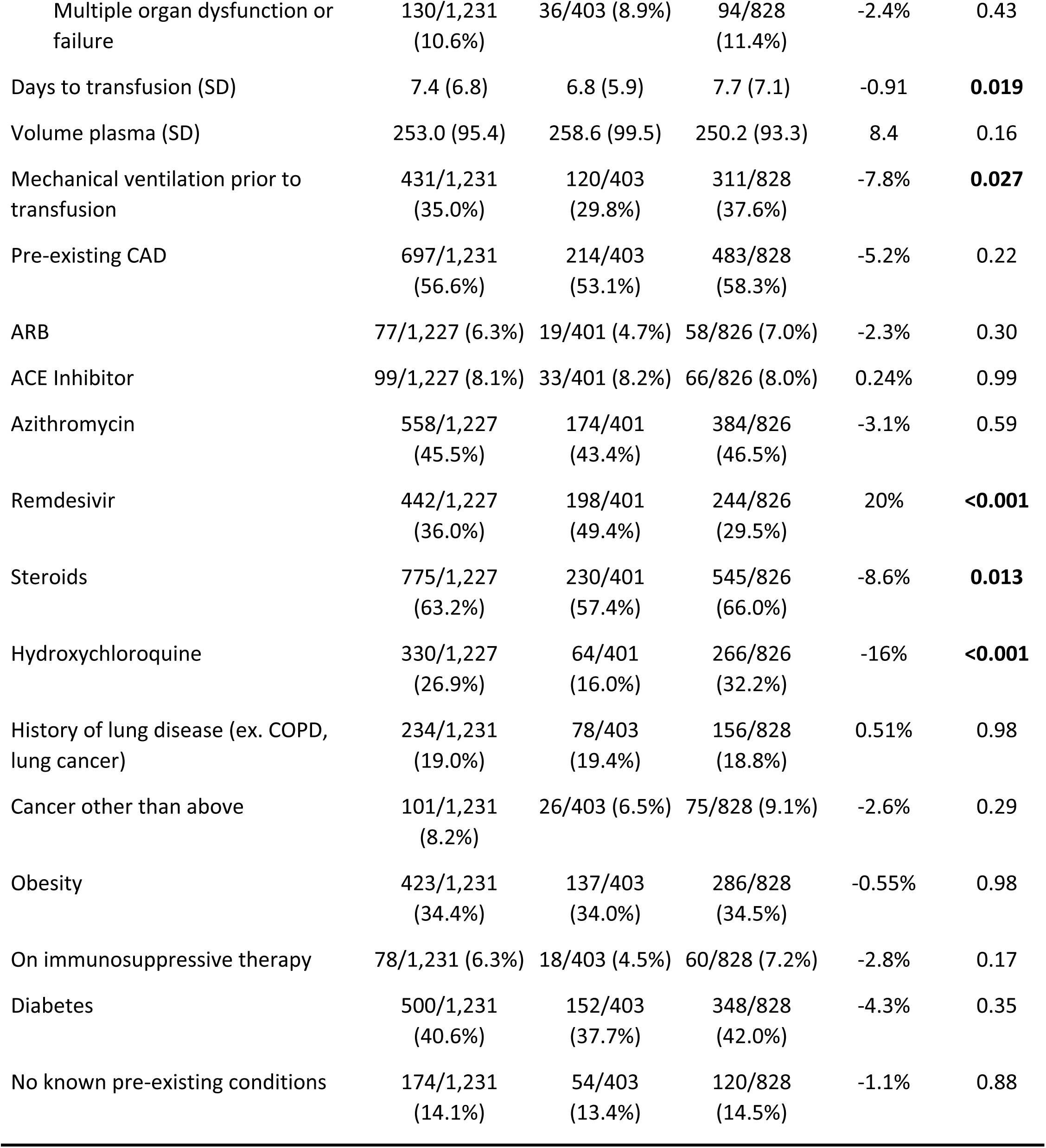
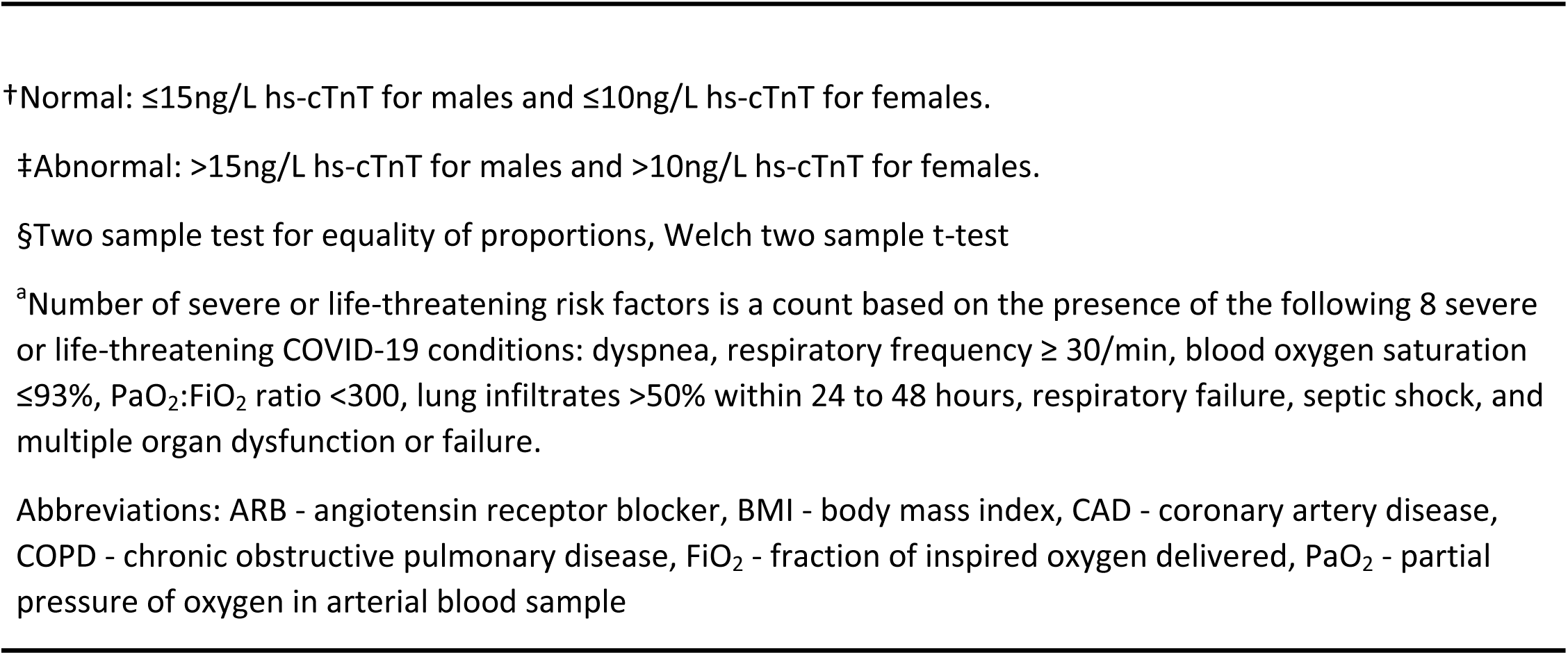
Epidemiology of patients with high/abnormal hs-cTnT levels.

### Troponin as a predictor of 30-day mortality using different cut-offs

Previous studies found that cTnI levels predicted mortality in patients with COVID-19;^2-4,6,9^ however, few studies examined hs-cTnT.^5,18^ In this study, we examined the effect of different widely accepted hs-cTnT cut-offs on mortality in severely ill patients that received convalescent plasma for COVID-19 (**Table 2**). When the FDA lowest allowable cut-off of <6ng/L was used, hs-cTnT levels did not predict mortality (Adjusted *P*=0.091). Using an ESC cut-off of 14ng/L,^16^ the odds of mortality were approximately two times higher in patients with elevated hs-cTnT compared to those with a normal level in both the unadjusted model and after adjusting for pre-existing cardiovascular disease (Unadjusted OR=2.06 [1.58 – 2.69], *P*<0.001, Adjusted OR=2.04 [1.56 – 2.67], *P*<0.001) (**Table 2**). Overall, adjusting for cardiovascular disease had very little effect on the data (**Table 2**). Using the sex-specific cut-off in males and females we found an increased 30-day mortality with elevated levels of hs-cTnT (mortality observed in 321 of 828 patients, 38.8%) compared to normal levels (mortality observed in 96 of 403 patients, 23.8%; *P*<0.001). The odds of mortality were around two times higher in patients with elevated hs-cTnT using the sex-specific cut-offs (Unadjusted OR=2.02 [1.55 – 2.66], *P*<0.001, Adjusted OR=2.00 [1.53 – 2.62], *P*<0.001) (**Table 2**), like our findings using the ESC cut-off of 14ng/L. The male-specific cut-off identified a 71% increase in the odds of mortality in patients with hs-cTnT >15ng/L compared to those ≤15 ng/L (Unadjusted OR=1.72 [1.24 – 2.41], *P*=0.001, Adjusted OR=1.71 [1.23 – 2.39], *P*=0.002). The female-specific cut-off identified a 169% increase in the odds of mortality in patients with hs-cTnT >10ng/L compared to those ≤10ng/L (Unadjusted OR=2.78 [1.76 – 4.52], *P*<0.001, Adjusted OR=2.69 [1.70 – 4.39], *P*<0.001). Among patients under the age of 65, those with an elevated hs-cTnT level (based on sex-specific cut-offs) had over twice the odds of mortality compared to those with a normal hs-cTnT level (Unadjusted OR=2.21 [1.47 – 3.39], *P*<0.001, Adjusted OR=2.2 [1.46-3.38], *P*<0.001). In patients 65 and older, elevated hs-cTnT levels were associated with 53% higher odds of mortality (Unadjusted OR=1.53 [1.06 – 2.22], *P*=0.024, Adjusted OR=1.53 [1.06 – 2.22], *P*=0.025). Values for hs-cTnT were divided into 4 groups by sex based on FDA or sex-specific cut-offs and shown as a percentage, lower and upper limits in **Figure 1**: ≤6ng/L, >6–15ng/L for males or >6–10ng/L for females, >15ng/L–99^th^ percentile for males, >10ng/L–99^th^ percentile for females; and >99^th^ percentile.^10,11,12^ There were no sex differences found in each of the groups (≤6ng/L *P*=0.2, >6ng/L-sex-specific cut-off *P*=0.76, sex-specific cut-off-99^th^ percentile *P*=0.12, and >99^th^ percentile *P*=0.56).

**Figure 1.**
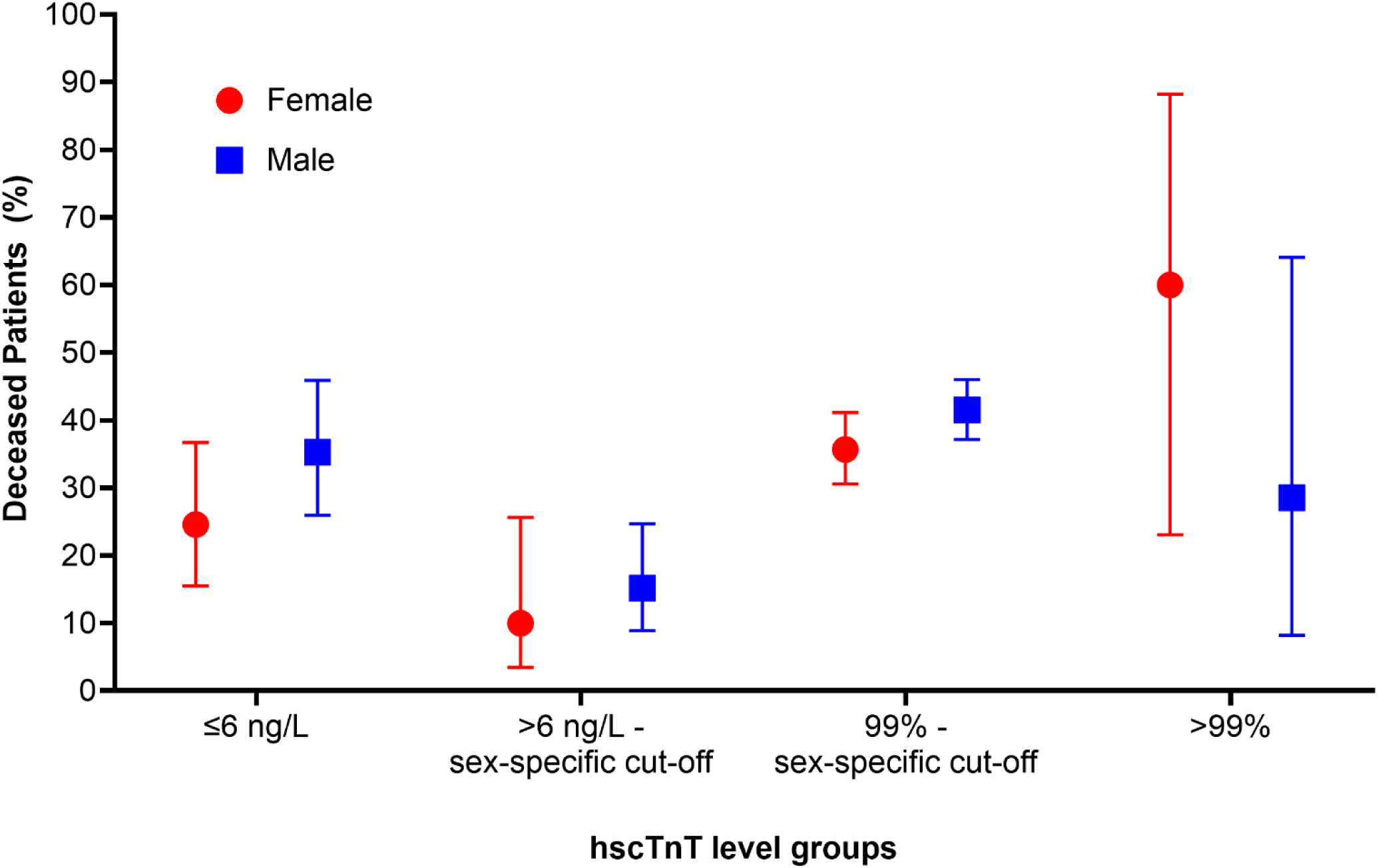
30-day mortality rates by hs-cTnT groups for males and females. The percentages are visualized with the lower and upper limits for each group. Sex-specific cut-offs are 10 ng/L for females (red) and 15 ng/L for males (blue). The 99^th^ percentile threshold is the ESC guidelines conjoint threshold.^16^

**Table 2:**
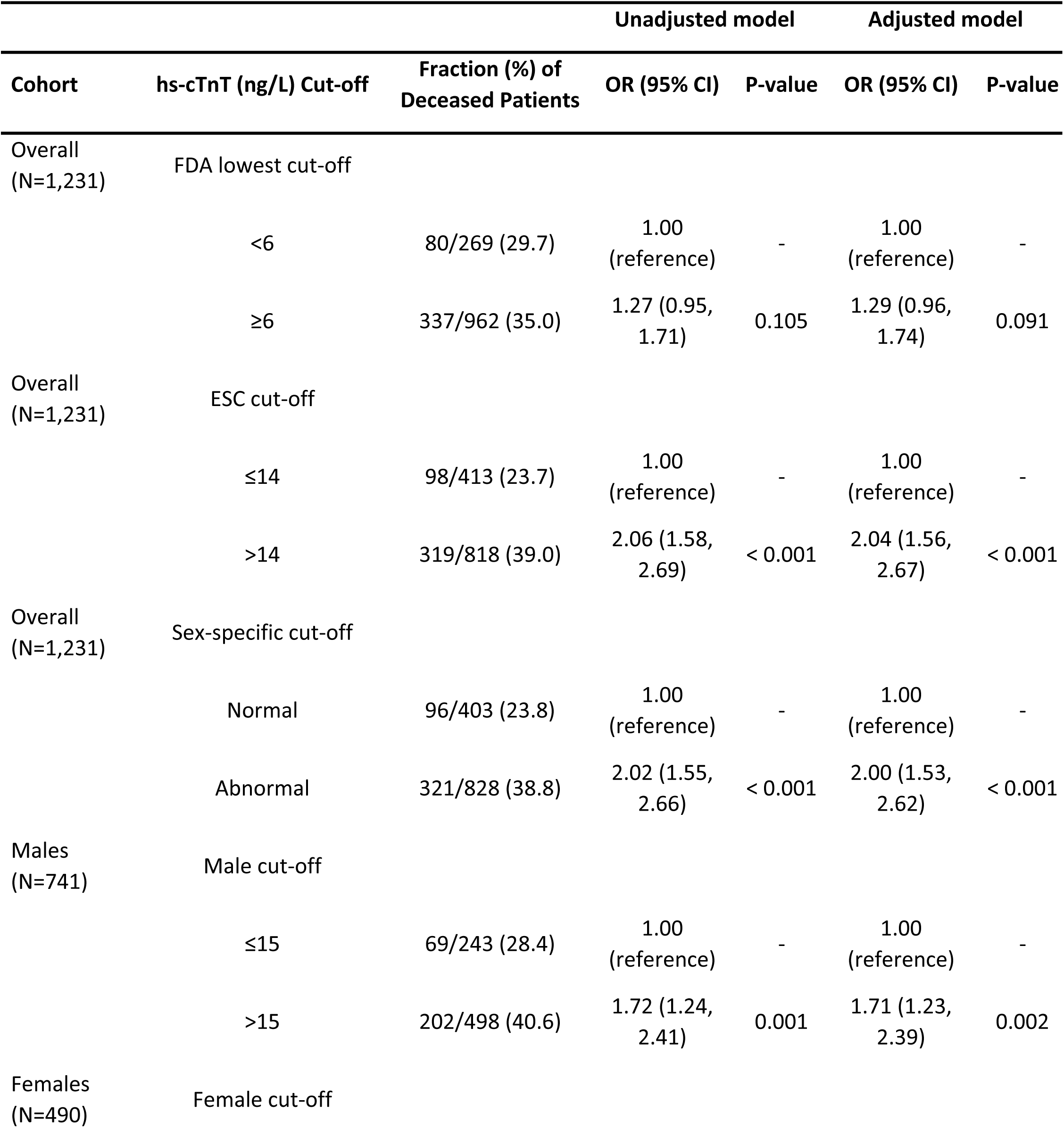
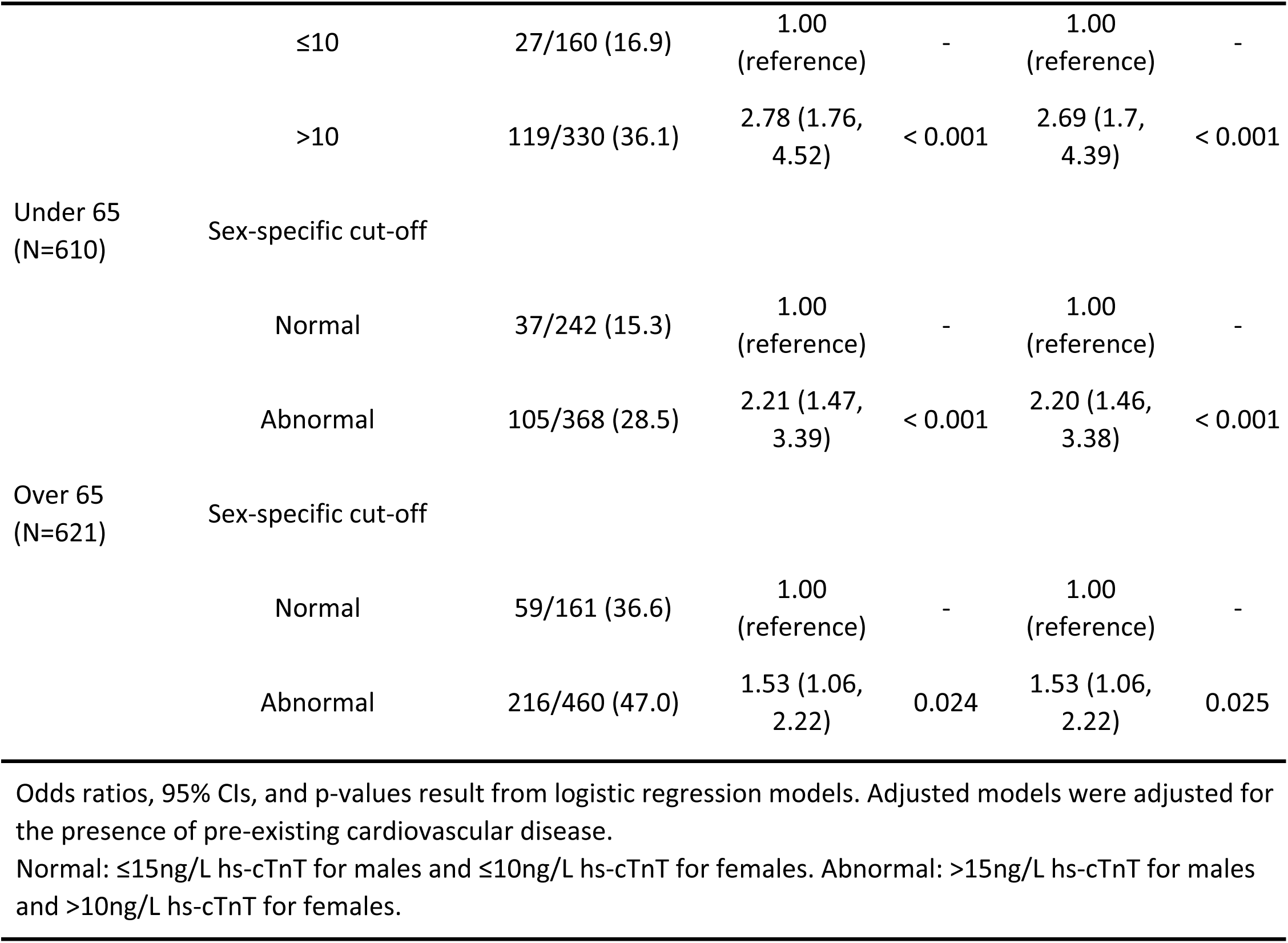
Troponin as a predictor of 30-day mortality using different cut-offs.

### Troponin as a continuous variable as a predictor of 30-day mortality

hs-cTnT levels significantly predicted 30-day mortality in the overall cohort (Unadjusted OR=1.18 [1.08 – 1.28], *P*<0.001, Adjusted OR=1.17 [1.08 – 1.28], *P*<0.001). This was also the case in the subset of male patients (Unadjusted OR=1.16 [1.05 – 1.29], *P*=0.005, Adjusted OR=1.16 [1.05 – 1.29], *P*=0.005) and female patients (Unadjusted OR=1.20 [1.04 – 1.39], *P*=.013, Adjusted OR=1.19 [1.03 – 1.37], *P*=0.019) or patients under 65 years of age (Unadjusted OR=1.19 [1.04 – 1.37], *P*=.013, Adjusted OR=1.19 [1.03 – 1.36], *P*=0.015) or over 65 years of age (Unadjusted OR=1.13, [1.02 – 1.26], *P*=0.026, Adjusted OR=1.13 [1.02 – 1.26], *P*=0.025) (**Table 3**). Overall, a 10-fold increase in hs-cTnT was associated with an 18% increase in the odds of mortality. In a separate model including a troponin-by-sex interaction term, no significant interaction was found (Unadjusted *P*=0.74; Adjusted *P*=0.79), indicating that the relationship between hs-cTnT and 30-day mortality as a continuous variable did not differ significantly between males and females.

**Table 3:** Troponin as a continuous predictor of 30-day mortality in various cohorts.

| Cohort | Unadjusted models |  | Adjusted models |  |
| --- | --- | --- | --- | --- |
|  | OR (95% CI) | P-value | OR (95% CI) | P-value |
| Overall (N=1,050) | 1.18 (1.08, 1.28) | < 0.001 | 1.17 (1.08, 1.28) | < 0.001 |
| Males (N=643) | 1.16 (1.05, 1.29) | 0.005 | 1.16 (1.05, 1.29) | 0.005 |
| Females (N=407) | 1.20 (1.04, 1.39) | 0.013 | 1.19 (1.03, 1.37) | 0.019 |
| Under 65 (N=498) | 1.19 (1.04, 1.37) | 0.013 | 1.19 (1.03, 1.36) | 0.015 |
| Over 65 (N=552) | 1.13 (1.02, 1.26) | 0.026 | 1.13 (1.02, 1.26) | 0.025 |

Odds ratios, 95% CIs, and p-values result from logistic regression models. Adjusted models were adjusted for the presence of pre-existing cardiovascular disease.
Note: 1,050/1,231 had continuous hs-cTnT (ng/L) data available. The predictor (hs-cTnT) was $\log_{10}$ transformed, meaning odds ratios represent the change in odds per 10-fold increase in troponin levels.

## Discussion

Our data adds significantly to the information available regarding the ability of cardiac troponin values to predict mortality associated with COVID-19. A number of studies found a significant relationship between cTnI levels at admission and adverse outcomes including mortality with COVID-19 early during the pandemic.^2-4,6,9^ Both hs-cTnI and hs-cTnT independently predict heart failure and mortality, but hs-cTnI may predict events in healthy individuals slightly better while hs-cTnT may be slightly stronger at risk prediction in older adults with comorbidities.^19,20^ Early during the pandemic, Cummings et al. found that most patients hospitalized with COVID-19 had elevated levels of hs-cTnT (*N*=257) indicating cardiac injury.^21^ Guo et al. demonstrated that COVID-19 patients with elevated plasma cTnT levels (52 of 187, 27.8%) were older, were more likely to be male, and had a higher prevalence of chronic cardiovascular conditions,^7^ similar to our findings in this study. Although the assay used was not described in the study, the values were in ng/ml suggesting this was not a high sensitivity assay. We previously showed that hs-cTnT levels were higher in men, were associated with more adverse events, and predicted 30-day mortality using sex-specific cut-off thresholds in hospitalized or emergency department patients with COVID-19 (*N*=367).^5^ These findings are similar to this study, but here we examined a larger number of patients (*N*=1,231) that were hospitalized for moderate to severe COVID-19 and we compared the prediction ability of several different widely accepted cut-off thresholds.

In this study we found that hs-cTnT values <6ng/L (i.e., the lowest reportable level) did not predict mortality, similar to previous findings.^5,18^ However, in our cohort, mortality, though lower in the low hs-cTnT group, was not nearly the zero percent as described in other studies.^2-9,18^ In this study using sex-specific cut-offs, we found that patients with normal hs-cTnT levels had 23.8% mortality. This likely reflects the severe or critical illness of patients enrolled in the EAP study and greater virulence associated with earlier strains of SARS-CoV-2. Nonetheless, 30-day mortality significantly differed between patients with normal and abnormal hs-cTnT levels (*P*<0.001). Furthermore, there was a graded relationship of hs-cTnT to mortality, with greater hs-cTnT levels associated with higher mortality.^18^ In this study, an 18% increase in the mortality rate was found for each 10-fold increase in hs-cTnT, which was reflected in the ability of hs-cTnT to significantly predict mortality when examined as a continuous variable. However, adjusting for prior cardiovascular disease had little effect on the ability of hs-cTnT to predict mortality as a cut-off or a continuous variable.

We found that the ESC cut-off of 14ng/mL and the sex-specific cut-offs for hs-cTnT when examined in males and females combined produced similar results (both *P*<0.001 with an OR of around 2). When examining data according to sex, we found that the OR using a sex-specific cut-off in males was 1.71 (*P*=0.002) but was 2.69 (*P*<0.001) in females using a female cut-off, suggesting that hs-cTnT may be a more sensitive predictor of mortality among females. However, using a troponin-by-sex model with continuous variables we did not find a sex difference in the ability of cTnT to determine mortality. Examining data by age, we found an OR of 2.20 for those under 65 years of age (*P*<0.001) and an OR of 1.53 for those over 65 years of age (*P*=0.025) suggesting that cTnT may be a better predictor of mortality in COVID-19 patients under the age of 65.

Cardiac troponin elevation, including both cTnT and cTnI, has been observed in patients with mild to moderate COVID-19, indicating myocardial damage. Several authors, such as Artico et al.,^22^ have proposed that myocardial injury in COVID-19 patients results from a combination of cytokine release, inflammation, and microvascular disease, although the exact mechanism remains unknown. According to Artico et al., patients with SARS-CoV-2 infection and troponin elevation due to myocardial injury have a significant relationship with cardiac abnormalities such as left and right ventricular impairment, pericardial effusions, and myocardial scarring. In these patients, myocardial injury appears to be related to macroangiopathy and microangiopathic thrombosis, which comes with the prothrombotic state of COVID-19. These data are similar to those reported by Metkus, who suggested that most injuries are common in all diseases that cause acute critical illness. Myocarditis and epicardial coronary artery occlusion leading to myocardial injury seem to be unusual.^23^

Nonetheless, as described by Bürgi et al.,^24^ even mild increases in troponin levels might be associated with a higher prevalence of cardiac complications in the future. This appears to be especially the case when myocardial scar is present. Scar after COVID-19 is a predictive marker of prognosis for future cardiovascular issues. It may be that those with more elevated hs-cTnT would have ventricular impairment and myocardial scar than those without elevated troponins. This may be important in the long term in distinguishing patients with a residual disease not due to COVID-19, such as patients with increases in cTn due to immunoglobulin complexes (macro troponins), leading to false positive results.^25^ These types of confounders are less frequently reported with hs-cTnT.^26^ Thus, when confirmed by imaging, the specificity of increases in cTn is significantly improved. Recent data suggests this phenomenon is common with some assays but less likely to occur with hs-cTnT.^26^

Limitations of our study include that we do not know whether the sex data provided in the case report forms is biological sex at birth or self-reported gender. Additional limitations of the study include that most of the patients in this study were critically ill, in contrast to most other studies which had a mix of low and high-risk patients. Thus, our data may only apply to this high-risk group. Additionally, all the patients in this study obtained convalescent plasma, which was associated with reduced mortality. ^12^ Additionally, the overall crude mortality in this cohort, where hs-cTnT values were available, was higher than the entire cohort in the EAP (∼34% vs. ∼25%),^12^ suggesting that sicker patients were more likely to obtain troponin testing. Finally, the troponin lab values utilized for this study could have been obtained from any point during a patient’s hospital stay. Thus, patients’ disease may have progressed, or treatment may have partially ameliorated it. hs-cTnT levels have been found to increase over time in COVID-19 patients, as has recently been reported.^18^ A statistical limitation of this study is the increased risk of Type I error due to the exploratory nature of the analysis and the evaluation of multiple models and associations without formal correction for multiple comparisons. Consequently, some observed associations may have occurred by chance.

In conclusion, our data indicate that increases in hs-cTnT are associated with increased mortality in COVID-19 patients hospitalized with severe disease. The magnitude of the effect varies by sex and age, suggesting that these covariates are important to consider in this population.

## Data Availability

The authors are not permitted to share the data that support the findings of this study directly. Individual participant data underlying the results reported in this publication, along with a data dictionary, may be made available to approved investigators for secondary analyses. Limited and de-identified data sets from the US COVID-19 Convalescent plasma EAP have been deposited into a research data repository and may be shared with investigators under controlled access procedures as approved by the Mayo Clinic Institutional Review Board. A scientific committee will review requests for the conduct of protocols approved or determined to be exempt by an Institutional Review Board. Requestors may be required to sign a data use agreement. Data sharing must be compliant with all applicable Mayo Clinic policies. Interested parties may contact the corresponding investigators. IRB approval will also be required for any access granted.

## Abbreviations

hs-cTnT: high-sensitivity cardiac troponin T
hs-cTnI: high-sensitivity cardiac troponin I
cTn: cardiac troponin
cTnT: Cardiac troponin T
cTnI: Cardiac troponin I
COVID-19: Coronavirus Disease 2019
EAP: US COVID Convalescent Plasma Expanded Access Program
CV: coefficient of variation
CVD: Cardiovascular Disease
LOD: limit of detection
LOQ: Limit of Quantification
OR: odds ratio
CI: confidence interval

## Acknowledgments

The authors thank the dedicated members of the US Convalescent Plasma Expanded Access Program team Machiko Anderson, Supriya Behl, Lori Bergstrom, Brian Butterfield, Grant Dubbels, Adam Eggert, Ree Erickson, Rebekah Frost, Daniel Gaz, Andrew Clayburn, Winston Guo, Riley Regimbal, Starr Guzman, Karina Hex, Vidhu Joshi, Megan Knudson, Tessa Kroeninger, Frances Lynch, Tim Miksch, Lisa Muenkel, Ryan Oldenburg, Amy Olofson, Laura Pacheco-Spann, Dr Kelly Avery, Dr Sumedha Penheiter, Melanie Peterson, Katrina Pierce, Nicloas Saikali, Jeffrey Schmoll, Pamela Skaran, Lindsay Stromback, Edward Swaray, Morgan Swope, Kristine Tree, Joe Wick, and Janelle Worthington; the members of the Mayo Clinic Institutional Review Board; the Mayo Clinic Office of Human Research Protection; the Mayo Clinic Office of Research Regulatory Support, and in particular Mark Wentworth. The authors thank the Executive Dean of Research at Mayo Clinic, Dr. Gregory Gores, and the CEO of Mayo Clinic, Dr. Gianrico Farrugia, for their support and assistance; the independent Data and Safety Monitoring Board for their work and oversight of the Expanded Access Program Drs Allan S. Jaffe (chair), David O. Warner, William G. Morice II, Paula J. Santrach, Robert L. Frye, Lawrence J Appel, and Taimur Sher; the members of the National COVID-19 Convalescent Plasma Project (http://ccpp19.org) for their intellectual contributions and support; the participating medical centers and medical teams and blood centers for their rigorous efforts necessary to make this program possible; the donors for providing COVID-19 convalescent plasma; Drs. Bruno and Jaffe are co-first authors. Drs. Bruno, Jaffe, Fairweather, Carter, Wright, and Joyner are co-senior authors.

